# An electronic application to improve management of infections in low-income neonatal units: pilot implementation of the NeoTree Beta App in a public sector hospital in Zimbabwe

**DOI:** 10.1101/2020.09.25.20201467

**Authors:** H Gannon, S Chimhuya, G Chimhini, S. R. Neal, L P Shaw, C Crehan, T Hull Bailey, R. A. Ferrand, N Klein, M Sharland, V Robertson, M Cortina Borja, M. Heys, F Fitzgerald

## Abstract

There are 2.9 million annual neonatal deaths worldwide. Simple, evidence-based interventions such as temperature control could prevent approximately two-thirds of these deaths. However, key problems in implementing these interventions are a lack of newborn-trained healthcare workers and a lack of data collection systems. NeoTree is a digital platform aiming to improve newborn care in low-resource settings through real-time data capture and feedback alongside education and data linkage. This project demonstrates proof of concept of the NeoTree as a real-time data capture tool replacing hand-written clinical paper notes over a 9-month period in a tertiary neonatal unit at Harare Central hospital, Zimbabwe. We aimed to deliver robust data for monthly mortality and morbidity meetings, and to improve turn-around time for blood culture results among other quality improvement indicators.

There were 3222 admissions and discharges entered using the NeoTree software with 41 junior doctors and 9 laboratory staff trained over the 9-month period. The NeoTree app was fully integrated into the department for all admission and discharge documentation and the monthly presentations became routine, informing local practice. An essential factor for this success was local buy-in and ownership at each stage of the project development, as was monthly data analysis and presentations allowing us to rapidly troubleshoot emerging issues. However, the laboratory arm of the project was negatively affected by nationwide economic upheaval. Our successes and challenges piloting this digital tool have provided key insights for effective future roll-out in Zimbabwe and other low-income healthcare settings.

## Problem

At Harare Central Hospital, Zimbabwe, approximately 12 000 babies are born each year and the 100-cot tertiary neonatal unit often runs at 140% capacity, admitting babies locally and nationwide for surgical management. Prior to this pilot quality improvement project documentation was paper-based and accessing records for audit and research purposes was both laborious and unrewarding, with excessive record loss. Quantification of even basic data such as admission and mortality rates was challenging, let alone measuring quality indicators such as temperature at admission. Junior doctors responsible for admitting babies spend 2 months on the unit before rotating, and senior support is overstretched. Junior doctors may have little experience of managing sick neonates, particularly identifying and managing sepsis acutely, and inappropriate antibiotics may be used. Nearly 60% of babies are admitted with presumed sepsis.[1] A recent Klebsiella sepsis outbreak had 33% case-fatality, and infection prevention and control interventions were hindered by delayed results from the laboratory and limited clinical documentation. Prior to this study, retrieval of microbiology results involved a doctor going in person to the laboratory, with a median turn-around time of 6 days. Delays in feedback of negative blood culture results likely prolonged admission and antibiotic therapy inappropriately while delays in positive culture results likely led to prolonged ineffective/excessively broad antimicrobial therapy, depending on sensitivities. In our baseline audit, 98% (449/459) of admitted babies received antibiotics at admission, and 99% (349/354) received oral amoxicillin at discharge, which is not an evidence-based intervention.[1]

NeoTree is a digital quality improvement platform co-developed with Malawian healthcare workers to improve newborn care in low-resource settings (see Crehan et al. for full description and screen grabs).[2] It offers education in newborn care, decision support, and suggested management plans according to country level and WHO guidelines alongside real-time data collection. The user-facing component of the platform is an Android application (app) on a tablet.

The aims of this study were to:

1. Demonstrate proof of concept as a real time data capture tool, replacing hand-written paper-based admission/discharge forms in the neonatal unit over a 9-month period
2. Deliver robust reliable data to be presented monthly at the neonatal unit morbidity and mortality meetings (within 6 months)
3. Improve the availability of data for diverse quality improvement projects such as antimicrobial stewardship and temperature control
4. Demonstrate proof of concept of NeoTree as a tool for surveillance of neonatal sepsis and antimicrobial use

## Background

Of 2.5 million annual neonatal deaths,[3] an estimated two thirds could be prevented through instigation of simple, evidence-based practices such as basic infection prevention and temperature control.[4] Efforts to reduce the rate of neonatal deaths are hampered by limited data collection, making it difficult to identify and prioritise modifiable risk factors for mortality. This in turn renders benchmarking and quality improvement measures challenging to evaluate and implement.[5]

Sepsis is implicated in ∼25% of neonatal deaths and many babies who do survive experience chronic morbidity.[6] Neonatal sepsis is characterised as early-onset (<72 hours of birth) or late-onset (>72 hours of birth), although these categories increasingly overlap, with both perinatal and healthcare-related risk factors contributing to each group.[6] In low-income settings overcrowding, understaffing, and restricted infrastructural and microbiological support render diagnosis and prevention of infections in neonatal units challenging, even in tertiary hospitals. A substantial shift to facility-based deliveries may have had the adverse consequence of rendering babies vulnerable from birth to bacteria typically associated with prolonged admissions (e.g. multi-drug resistant Gram-negative organisms may cause sepsis in the first 24 hours of life).[7]

Pilot work in Malawi using and iteratively developing the NeoTree alpha version suggested a high degree of user satisfaction with NeoTree, with feedback used to upgrade the platform to a Beta version; Minimal Viable Product (MVP-1). NeoTree has the potential to streamline record keeping, feed-back results and contribute to improved sepsis surveillance, as well as providing guidance to clinicians and healthcare workers about management of key neonatal diagnoses. We piloted the Beta version (MVP-1) of the NeoTree at Harare Central Hospital neonatal unit and additionally developed the lab data collection pages. We hypothesised that a novel app page for feeding back blood culture results from the laboratory to the neonatal unit could reduce test turn-around times. Establishing NeoTree as a robust data collection platform and a tool for antimicrobial surveillance could be a first step in instigating national level surveillance of antimicrobial resistance in neonatal units.

## Measurement

Our setting was Harare Central Hospital neonatal unit, with the population being all admitted neonates over a 9-month pilot period (November 2018-July 2019).

Prior to implementation of NeoTree, a prospective audit was carried out over a month period to measure baseline admission/discharge rates and case fatality rates, antibiotic prescription rates and blood culture results and turn-around time.[1] The previous standard of documentation to collect total numbers of admissions/discharges and deaths was a hand-written book, held and completed by the sister-in-charge. There were 459 admissions over 28 days with an overall case fatality rate of 210 per 1000. Blood culture results were fed back in a median of 6 days, with 7/196 (3.6%) cultures turned around in time for clinical utility; i.e. in time to impact on therapy. Oral amoxicillin at discharge was prescribed for nearly all babies despite a lack of evidence for efficacy, though this dropped dramatically to 1/161 babies (0.6%) at repeat audit with intensive education and training for junior doctors prior to NeoTree introduction.

Our aim for this quality improvement project was to demonstrate proof of concept of NeoTree as a real-time data capture tool, replacing hand-written paper notes over the 9-month period and to be able to provide monthly results to the neonatal unit. The primary end point measurement was to measure the number of admissions, discharges and deaths captured on the NeoTree when compared to the current standard of documentation within the unit; the admission/discharge/death hand-written book. Our target was for 100% of admissions, discharges and deaths to be recorded on the NeoTree app at 9 months. Blood culture results turn-around time was to be measured and compared to the baseline data result.

To measure the ability to provide monthly data to the neonatal unit staff by month 6, we targeted month 3 to commence monthly meetings, taking a register of attendance and implement a culture of learning and feedback within these sessions. The sessions were to involve a register taken, creation and delivery of a powerpoint presentation of the data monthly and feedback regarding the usability of the NeoTree itself with suggestions and improvements, documented and instigated as soon as possible.

For data collection during the project, NeoTree-Beta acted as a real-time data collection tool, with pseudonymised data from each admission and discharge form being stored on the tablet after a hard, patient identifiable copy was printed for the notes (currently only paper-based notes have legal standing in Zimbabwe). These pseudonymised data were exported daily to a secure server where data were collated and analysed using R version 3.6.0 (R Core Team, Vienna, Austria) with RStudio version 1.2.1335 (RStudio Team, Boston, United States)[8], on a monthly basis. For the laboratory page (NeoLab), laboratory staff would enter blood culture results onto the laboratory tablet when available including preliminary negative results at 48 hours. These results were printed immediately on the neonatal unit via a WiFi connection where the junior doctors could collect them and file them in patient notes. Time of filing in notes was collected.

## Design

The team for the project consisted of Zimbabwean and UK members. UK members included clinicians, software developers and statisticians, while Zimbabwean team members were consultant clinicians at Harare Central Hospital. UK members were responsible for ongoing software development in response to clinician feedback, data management and overall project logistics. Prior to instigation a round of informal usability testing was completed. This involved local staff completing an admission and discharge form using the app, giving feedback as they progressed. This feedback informed country and facility specific adjustments to the Malawi NeoTree MVP-1 admission and discharge forms to produce ‘Zimbabwe MVP-2’. The format of the NeoTree admission/discharge pages were altered using an editor platform which requires minimal software expertise to use (i.e. without having to consult the software team) to account for specific unit needs. These included how the platform would fit within current patient flow. Zimbabwean team members tailored the clinical management pages to ensure local relevance.

We hired a staff member (the ‘NeoTree Ambassador’) responsible for checking tablets into and out of the secure locker where they were stored, charging tablets, data export, day-to-day supervision of the junior doctors to ensure the forms were correctly filled out, checking all babies admitted/discharged were being entered onto the NeoTree, training new staff members and acting as a project advocate within the unit (e.g. explaining the project to families).

Based on previous experience in Malawi,[2] we planned to implement the Zimbabwe NeoTree MVP-2 gradually over 4 weeks, initially with a few admissions/discharges with each junior doctor per day supported by the study coordinator/Ambassador, then unsupervised day time admissions/discharges, followed by weekends and then night time shifts until all admissions/discharges would be captured. Each individual was trained by the study coordinator/NeoTree Ambassador prior to being allocated a tablet both in the unit and the laboratory. Discharge and laboratory forms were to be matched with admission forms using a unique identifier (NeoTree number) generated by the app on admission as data stored on the server were pseudonymised. Prior to implementation, we carried out a series of sensitisation training sessions for nurses, healthcare assistants, cleaners and administrative staff to ensure buy-in and enable all staff to answer questions that parents or family members might have. We held a monthly feedback session during the unit weekly meeting to present key indicator data from the previous month, and to encourage suggestions from staff about both potential improvements to the app and quality improvement questions that the data could be used for. Small monthly cash prizes were to be awarded to the junior doctor producing the most accurate ‘NeoTrees’ each month (i.e. the number of admission and discharge/death forms accurately completed). Suggestions for app improvements were also encouraged on a day-to-day basis from junior doctors, and where feasible these were rapidly incorporated within the app scripts to encourage local ownership and satisfaction, usually by the clinician project coordinator using the editor platform. It was rarely necessary to involve the software team in edits.

Ensuring the application was locally relevant and responsive was a key strategy for future-proofing the app. We were keen to make the app as usable and useful as possible for the junior doctors, and to encourage locally led ideas for quality improvement. Another part of planning for future sustainability was engaging with the Zimbabwean Ministry of Health and Child Care who had an established Electronic Medical Records Department. They had not yet developed a neonatal ‘module’, and so were keen to discuss how NeoTree could be embedded within local systems for future wider roll out.

## Strategy

### LEADERSHIP AND CO-DEVELOPMENT

Our strategy for roll out depended heavily on leadership within the unit. The medical and nursing senior team within the department were enthusiastic about the platform’s potential to improve care and data collection and provided vital support and encouragement to the junior doctors using the tablets. They also provided a real-time system of quality control: if there were inaccuracies or omissions on the admission form, these were picked up by seniors during daily ward rounds and fed back to the juniors. Similarly, discharge summaries were reviewed both in follow-up clinics and in spot checks on the unit by senior clinicians. The project coordinator directly canvassed opinions from junior doctors/nurses about potential improvements and problems, initially on a daily, then weekly, then monthly basis, although the NeoTree Ambassador was available every day to answer queries, troubleshoot and collect suggestions from frontline staff.

### IMPLEMENTATION LESSONS

The monthly data feedback sessions acted as host for our progressive improvement cycles as described below. From the laboratory side, there were considerable hurdles with availability of culture media and then with staffing issues (see ‘lessons learned’ section). Industrial action also impacted the neonatal unit itself. We undertook 4 separate Plan-Do-Study-Act (PDSA) cycles during the 9-month period.

### PDSA 1: Revision of death discharge forms

In month 2, we noted incomplete capture of deaths on the discharge/death forms. This was when the number of deaths on the NeoTree were compared to the numbers of deaths documented in the hand-written death/discharge book recorded by the sister in charge. It came to light that from the 154 deaths documented from months 1 to 3, 143 (93%) had been completed by the NeoTree ambassador and not the healthcare workers using the NeoTree. Feedback from juniors highlighted that where the admission/discharge electronic form replaced the paper forms, the NeoTree death forms were duplicates of effort as statutory reporting of deaths mandated that deaths were documented on special government forms that could not be replaced. We addressed this by intensifying scrutiny by the NeoTree staff to ensure all babies who had died had been captured within the app, liaising directly with juniors and emphasising the importance of the data entry (with support of senior clinical staff). We also altered the monthly prize to incentivise the input of deaths onto the NeoTree. Over the next two months the capture of deaths improved, although still needing ongoing input from NeoTree staff. Unfortunately, at month 7 it was found that the number of deaths documented had again decreased and particularly babies dying very shortly after admission were not being captured. We instigated a rigorous audit programme with support of senior clinical staff and allowed a shortened discharge/death form to be completed without a separate admission form to be completed for these babies. Supplementary Figure 1 demonstrates the trend of death documentation throughout the project.

During the course of the study we commenced discussions with senior hospital management and the Ministry of Health to allow a NeoTree printout to be acceptable as formal death documentation. We believe while there is still duplication of effort, death documentation will be a weakness of NeoTree needing continuous monitoring and team input.

### PDSA 2: Revision of NeoTree ID number

By month 3 we found a high number of ‘unmatched’ discharge forms where the NeoTree ID entered on the discharge form had no corresponding NeoTree ID on an admission form. This was partly due to admission forms not being completed if the baby died shortly after admission (as mentioned under PDSA 1), but the percentage of unmatched discharge forms exceeded the level that would be expected if this were the sole cause. It was apparent that NeoTree ID numbers were frequently entered incorrectly on the discharge form. We addressed this partly via software changes using ongoing version control (software code available at https://github.com/neotree/analytics) – shortening the ID number to 8 digits and using ‘fuzzy logic’ to match admissions/discharges based on common mismatches in the dataset where e.g. 0 (the number) and O (the letter) had been confused – and also via feedback to junior staff with a further amendment of the monthly prizes to include a ‘league table’ of highest percentage matches between the junior doctors. This successfully reduced the number of NeoTree ID mismatches, dropping from 44% unmatched to <10% unmatched (Figure 1).

**Figure 1.**
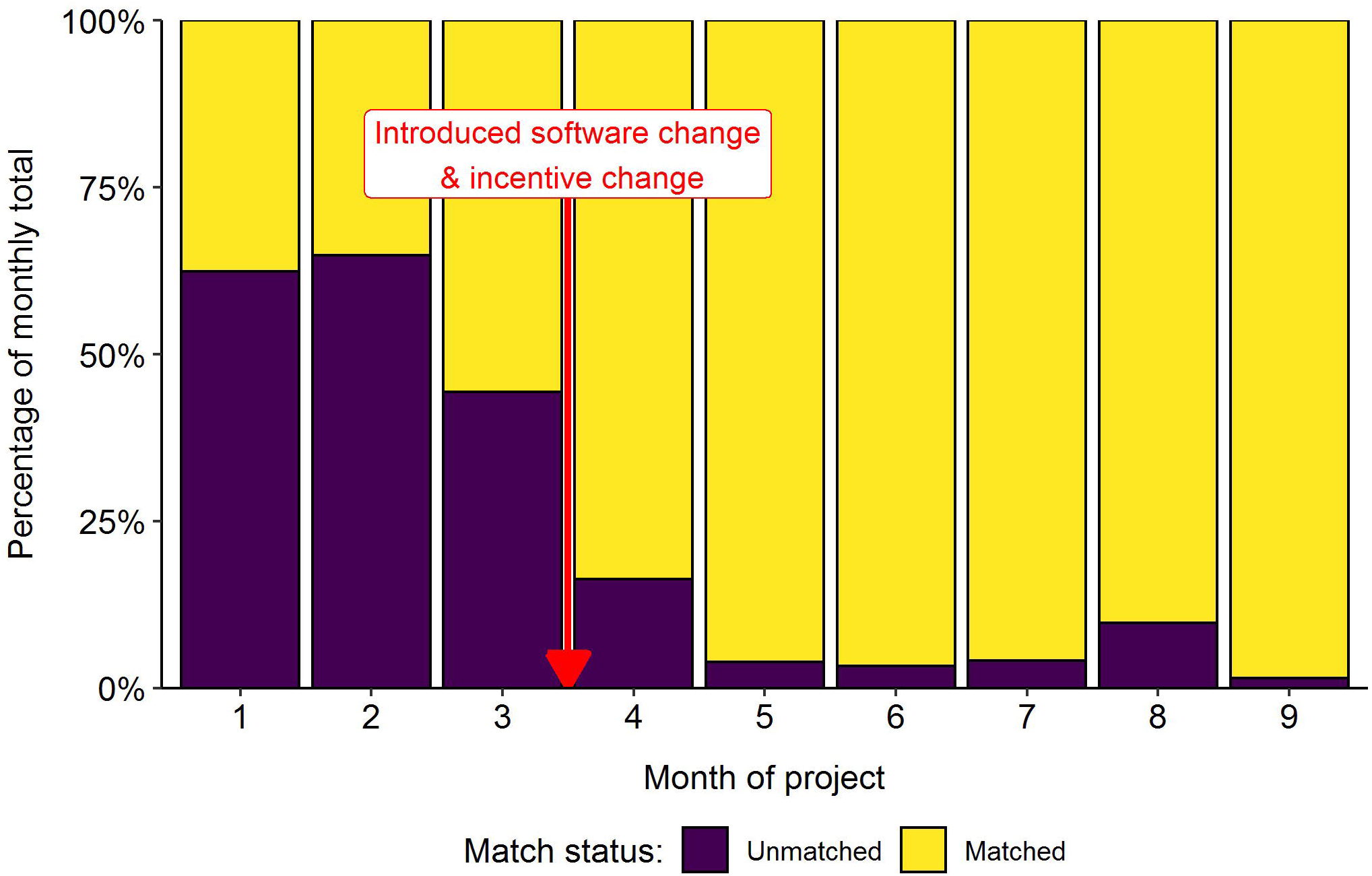
Trend in NeoTree ID matches per month throughout the project.

In a current parallel project, we are investigating the use of record linkage techniques, including probabilistic record linkage,[9] to improve record matching and to increase the proportion of matched admission and discharge files. The next iteration of the NeoTree app has inbuilt functionality to match ID numbers with currently admitted patients at discharge to minimise mismatch risk.

### PDSA 3: Antibiotic prescription rates at discharge

Each month we reviewed key statistics as suggested by medical/nursing staff. For example, in month 5, it was requested that we review amoxicillin prescription at discharge and found the figures to have increased again to 78/354 (22%) of discharged babies since the previous audit. We undertook further training for doctors as well as nursing staff in antimicrobial stewardship and the lack of rationale for amoxicillin use. This training was carried out both informally on ward rounds and formally during weekly unit teaching sessions and included amoxicillin statistics in the regular feedback at the monthly meeting. The rate of amoxicillin prescription reduced to 8/359 (2%) the following month and has remained low, although scrutiny is ongoing (Supplementary Figure 2).

### PDSA 4: Thermoregulation data

Hypothermia at admission was another key indicator. Initially, this indicator was often missing. Temperatures were routinely taken by nursing staff on admission to the unit rather than junior doctors on first review of the baby (e.g. in the labour ward) so this was often left blank. Less than 40% of babies in month 2 and 3 had their temperatures recorded. We supplied thermometers for the two doctors on call and made the data entry field mandatory as opposed to optional, which improved the documentation. After this implementation the percentage of babies with temperatures recorded increased and, by month eight, had increased to 90% (Figure 2).

**Figure 2.**
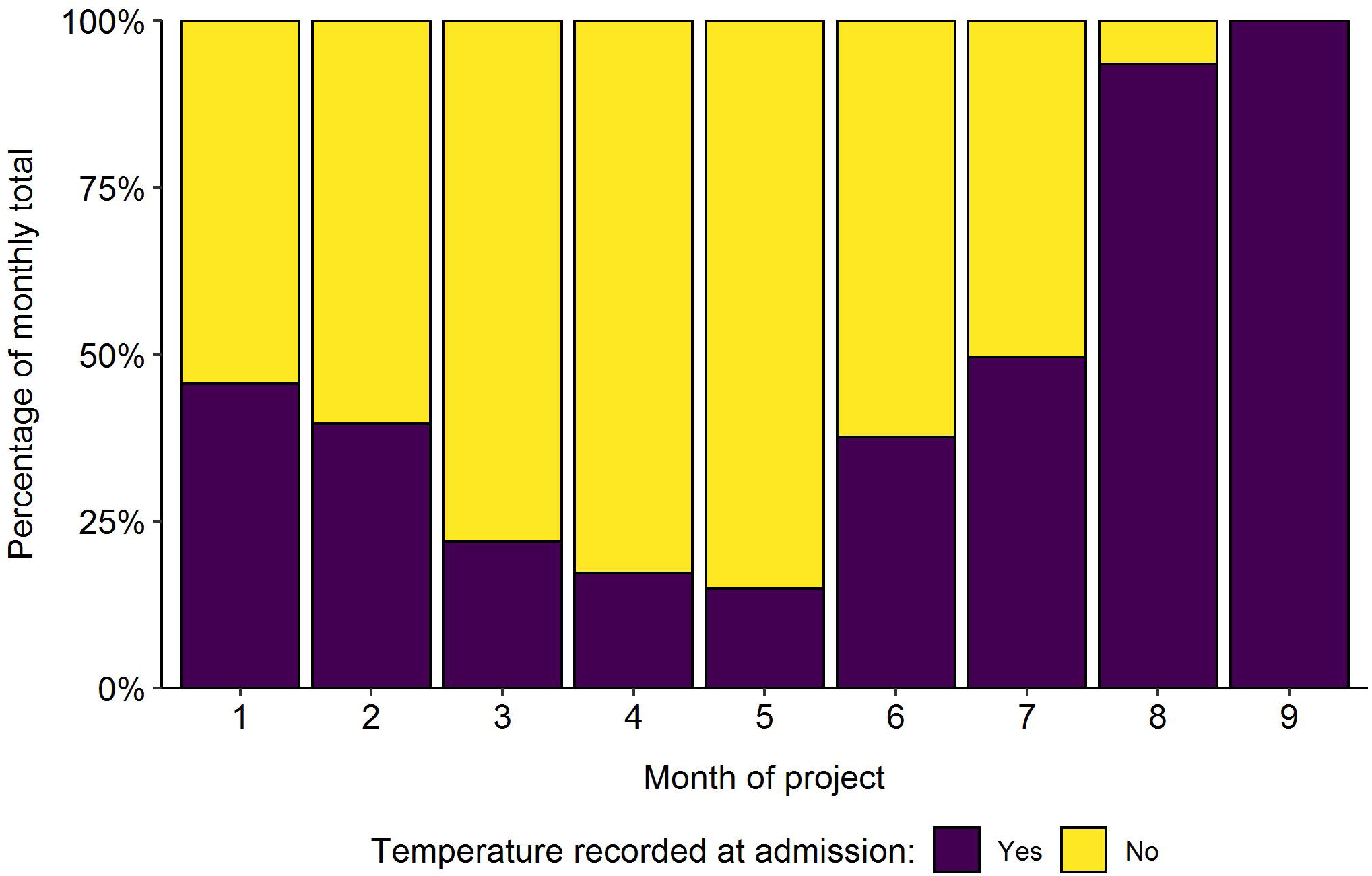
Percentage of babies with their temperature measured at admission throughout the project.

From discussions with unit staff, it was felt that most babies admitted with hypothermia were outborn rather than inborn. However, we showed that the majority of babies who were hypothermic were actually inborn, and to instigate a programme of ensuring that small, premature babies were supported with adequate temperature control (Supplementary Figure 3). This is an ongoing project as we have not yet seen an improvement in temperatures at admission (Supplementary Figure 4). We believe this is partly due to it being winter in Zimbabwe at the time of the PDSA cycle commencing (meaning ambient temperatures can be as low as 5°C at night) and partly to do with the doctor’s strike.

The project was approved by Harare Central Hospital Ethics Committee (HCHEC 250418/48) and by University College London Ethics Committee (5019/004). As this was a quality improvement project aimed at strengthening routine clinical care and data were pseudonymised, requirement for individual consent was waived.

## Results

We trained 41 junior doctors, 9 laboratory staff and sensitised 94 nursing/midwifery staff. We presented preliminary audit data both at a national level and to the hospital executive team. There were 3,222 admissions and discharges/deaths of babies entered using the NeoTree software, despite a 6-week doctor strike and a national level shut down including government shutdown of all internet across the country. Monthly admission, discharge and mortality data are shown in Figure 3. We fulfilled both aims 1 and 2 by providing admission, discharge and death data on a monthly basis for the unit including the basic statistics required for hospital management feedback such as mortality broken down into term/preterm, mortality by birth weight, causes of death, receipt of prevention of mother to child transmission therapy for HIV and admission/discharge diagnoses (Supplementary Table 1). We also provided data for locally led QI projects such as antimicrobial stewardship which has resulted in a sustained decrease of unnecessary prescriptions of oral amoxicillin at discharge (Supplementary Figure 2). Adherence to appropriate first line antimicrobial therapy has improved: at baseline, 9% of babies received ceftriaxone as opposed to crystalline penicillin and gentamicin. At last review 1.5% of babies received ceftriaxone as first line therapy. The hypothermia QI project is ongoing.

**Figure 3.**
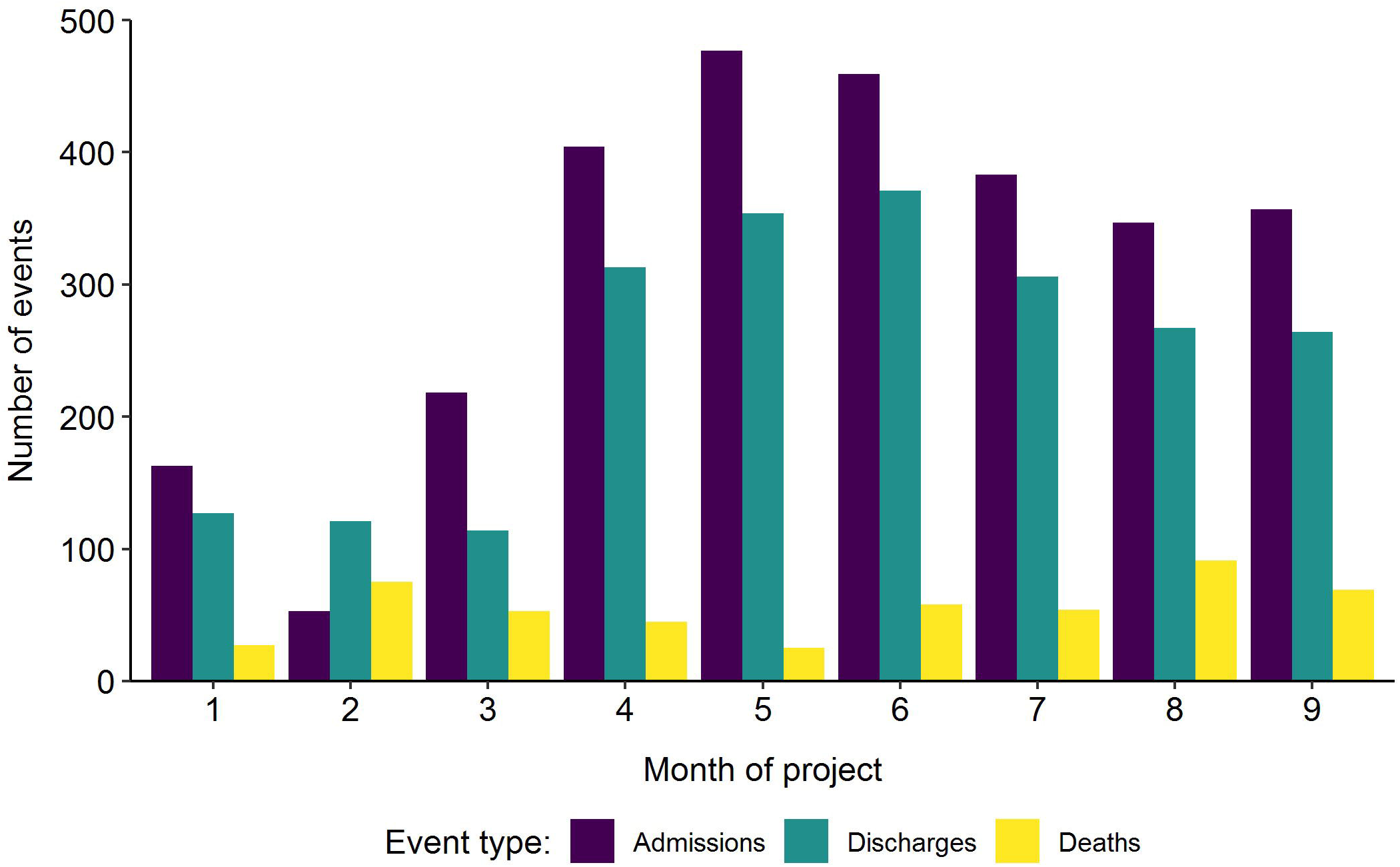
Frequencies of admissions, discharges and deaths per month throughout the project.

However, the laboratory arm of the project was and continues to be more challenging. Initially, we reduced laboratory turn-around time for blood culture results from 6 days to 3 days within the first 6 weeks of introduction. However, this was not sustained (see ‘Lessons and limitations’ below), and the current turn-around time for results to be fed back using the NeoTree is between 6 to 10 days. We have unfortunately also had such incomplete laboratory data that we have not yet been able to develop the surveillance platform.

The NeoTree data are currently being used for two further locally led quality improvement projects: management of congenital syphilis and management of late preterm infants. In addition, NeoTree data will be used in Zimbabwean-led research projects in neonatal sepsis, antenatal steroid use and hypothermic ischaemic encephalopathy (this last project in conjunction with developing a similar platform for mothers in labour with Zimbabwean obstetricians - the ‘MummyTree’). We are in advanced discussions with the Ministry of Health about how to incorporate the NeoTree into their plans for nationwide electronic medical record roll out, and with the Medicine Control Authority of Zimbabwe about how the NeoTree can be used to track birth defects associated with antiretroviral therapy for HIV.

## Lessons and limitations

The most challenging aspect of the project was the laboratory side. After an initial 6 weeks where we improved turn-around time, there were then five months of issues with media availability, meaning no cultures were performed. When the media finally became available again, economic upheaval and industrial action led to a policy of ‘flexible working’ in the laboratory, meaning skeleton staffing became the norm. Despite financial incentives from the NeoTree project morale was very low, and it was increasingly challenging to motivate staff to complete the laboratory form. Further interventions are ongoing, but this aspect of the project has suffered from force majeure. By contrast, despite industrial action by junior doctors in the second and third month of the project, the remaining skeleton junior staff (two out of a rostered eleven) were strong advocates for NeoTree and continued to use it although more for discharges than admissions. This meant when the full staffing complement returned in January 2019, using NeoTree was the departmental norm, and there was considerable peer-to-peer training in addition to that provided by NeoTree staff. In general, the junior doctors were familiar with touch-screen technology and quick to learn the process of using the app and printing the forms. Availability of reliable WiFi provision (needed for connecting tablets to a printer and exporting data) and power was key. These facilitating factors may not be reproducible in other settings, particularly power and the staff cadres using NeoTree. The app is designed to work offline with data exported intermittently, maintaining the education functionality, although alternative printing arrangements such as Bluetooth options would need to be in place. A variety of staff cadres found NeoTree to be highly ‘usable’ in Malawi, albeit with more training and practice.[2]

## Conclusion

We have shown the NeoTree app to be an effective tool for data capture, replacing hand-written paper-based admission, discharge and laboratory forms within HCH neonatal unit. The data captured were routinely fed back to the unit during monthly presentations, when regular feedback was taken about the app, with subsequent iterative improvements made and further locally driven quality improvement projects commenced. These data were presented at hospital executive and national level, to guide future management within the unit. Antimicrobial stewardship was supported by effective surveillance of amoxicillin at discharge. However, despite the successful integration of the NeoTree into the neonatal unit, the laboratory arm suffered from challenges often encountered in low-income settings, namely economic upheaval, industrial action and shortages of supplies. We are continuing to work towards our aim of implementing a sepsis surveillance platform.

The NeoTree app has been embedded into usual clinical practice for admission and discharge documentation, with the monthly presentations now normal practice within the unit, guiding and changing local practice with minimal external input from the NeoTree team. An essential factor for this success was strong local leadership. Regular feedback and the ability to adapt to local needs is a vital attribute of the NeoTree project. The next steps are planned piloting in a provincial hospital to test usability in a nurse-led unit using the iterative PDSA processes as described above. Economic analysis (currently ongoing) will be crucial to ensure feasible further roll-out as well as ensuring the platform is robust in a wider variety of settings.

## Data Availability

All data relevant to the study are included in the article, uploaded as supplementary information
or available upon request from the corresponding author.

## Acknowledgements

We are grateful to the staff, infants and their families at Harare Central Hospital, Dr Christopher Pasi and Matrons Alice Mudzingwa and Dade Pedzisai for their support.

## SUPPLEMENTARY FIGURE LEGENDS

**Supplementary Figure 1.**
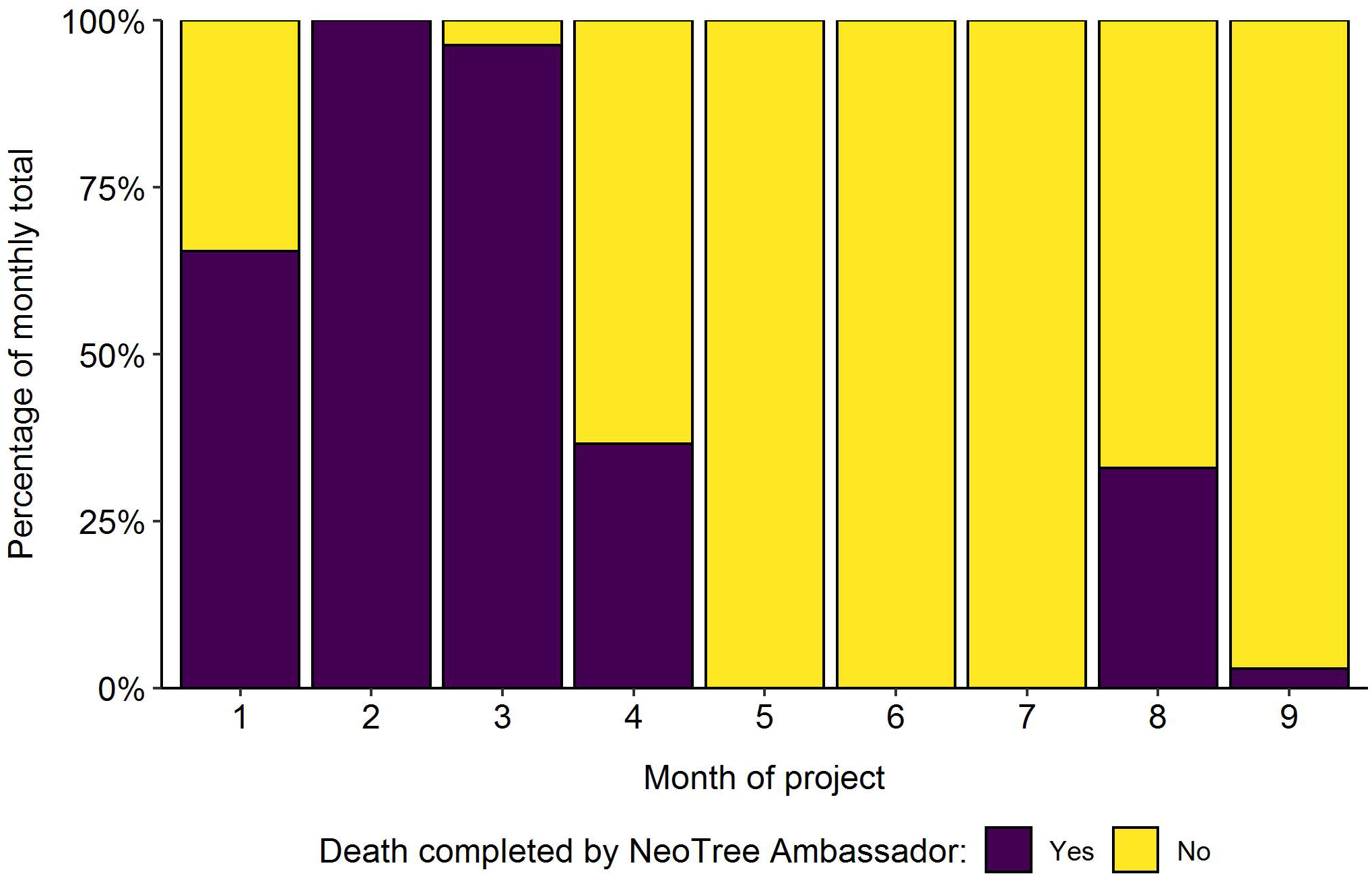
Percentage of deaths completed by the NeoTree Ambassador per month throughout the project.

**Supplementary Figure 2.**
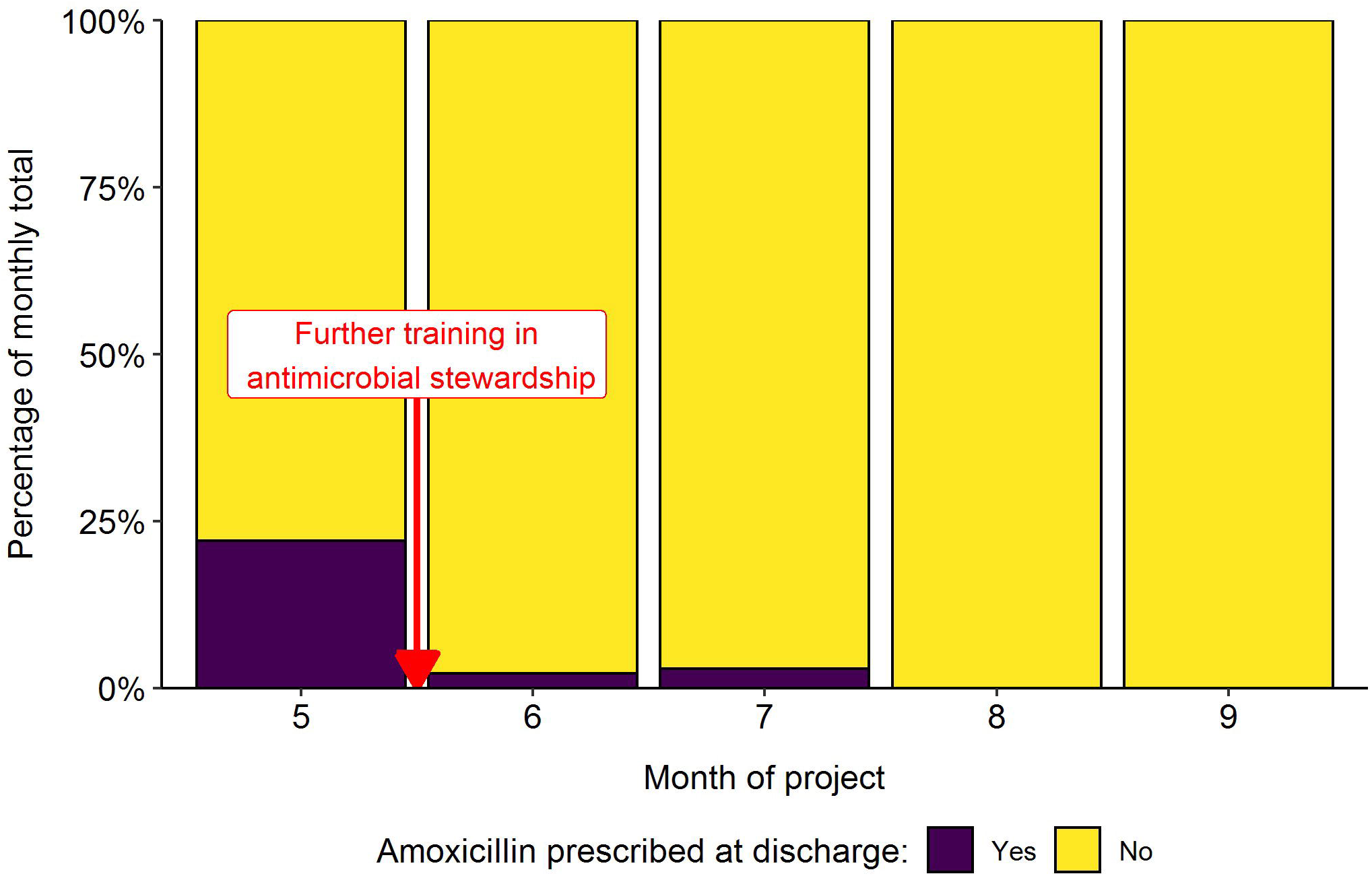
Trend in amoxicillin prescriptions at discharge per month from month 5 of the project

**Supplementary Figure 3.**
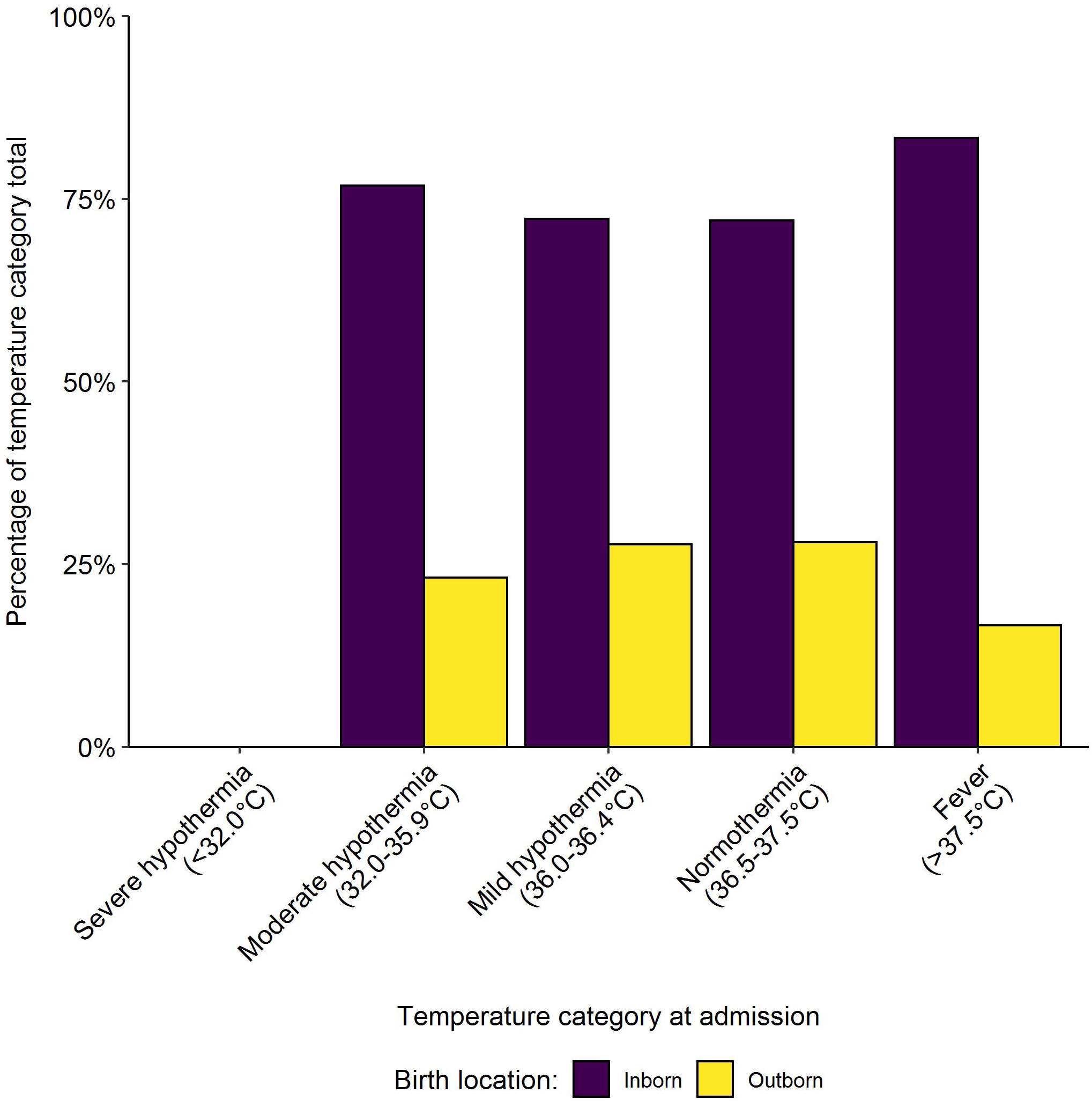
Percentage of babies in each temperature category at admission by birth location (inborn or outborn) for month 8 of the project.

**Supplementary Figure 4.**
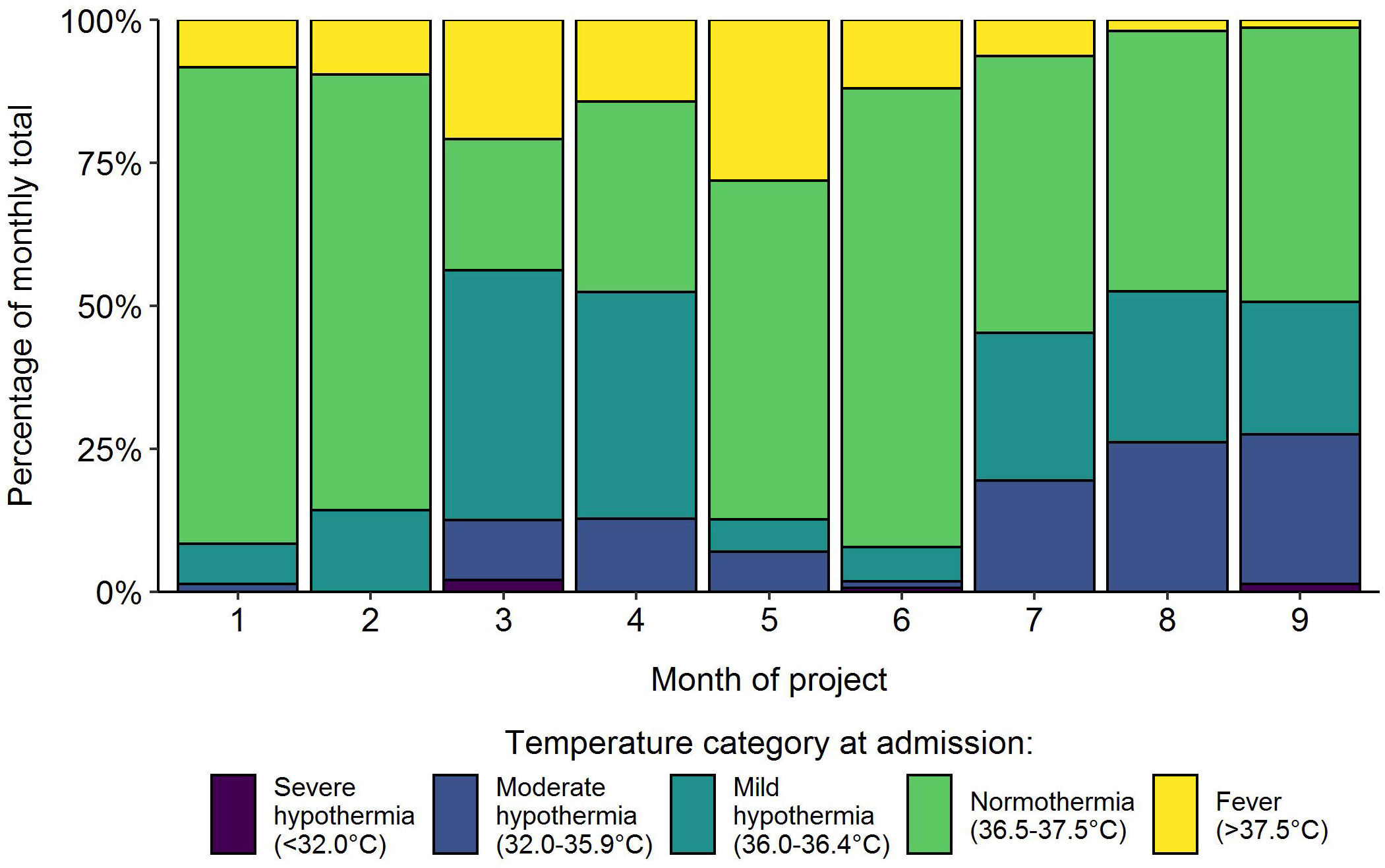
Trend in temperature category at admission per month throughout the project. Severe hypothermia: <32°C Moderate hypothermia: 32.0-35.9°C Mild hypothermia:36.0-36.4°C Normothermia: 36.5-37.5°C Fever: >37.5°C

## References

[1] Chimhini G, Chimhuya S, Madzudzo L, et al. Auditing use of Antibiotics in Zimbabwean neonates. Infection Prevention in Practice June 2020;

[2] Crehan C, Kesler E, Nambiar B et al. The NeoTree application: developing an integrated mHealth solution to improve quality of newborn care and survival in a district hospital in Malawi. BMJ Glob Health 2019; 4: e000860.

[3] United Nations Inter-agency Group for Child Mortality Estimation. Levels & Trends in Child Mortality: Report 2019. New York: United Nations Children’s Fund; 2019.

[4] Knippenberg R, Lawn JE, Darmstadt GL et al. Systematic scaling up of neonatal care in countries. Lancet 2005; 365: 1087–98.

[5] Lawn JE, Blencowe H, Oza S et al. Every Newborn: progress, priorities, and potential beyond survival. Lancet 2014; 384: 189–205.

[6] Fitchett EJA, Seale AC, Vergnano S et al. Strengthening the Reporting of Observational Studies in Epidemiology for Newborn Infection (STROBE-NI): an extension of the STROBE statement for neonatal infection research. Lancet Infect Dis 2016; 16: e202–e13.

[7] Zaidi AKM, Huskins WC, Thaver D, et al. Hospital-acquired neonatal infections in developing countries. Lancet 2005; 365: 1175–88.

[8] Team RC. R. A language and environment for statistical computing R Foundation for Statistical Computing, Vienna, Austria (2017) (Version 3.6. 0)[Computer software]

[9] Herzog TN, Scheuren FJ, and Winkler WE. 2007. Data Quality and Record Linkage Techniques. 1st ed. Springer-Verlag, New York. ISBN: 978-0-387-69502-0. DOI: 10.1007/0-387-69505-2.

